# Increased T2 Relaxometry in Mild Traumatic Brain Injury: An Individualised Marker of Acute Neuroinflammation?

**DOI:** 10.1101/2024.03.10.24303890

**Authors:** Mayan J. Bedggood, Christi A. Essex, Alice Theadom, Samantha J. Holdsworth, Richard L.M. Faull, Mangor Pedersen

## Abstract

Mild traumatic brain injury (mTBI), often called concussion, is a prevalent condition that can have significant implications for people’s health, functioning and well-being. Current clinical practice relies on self-reported symptoms to inform return to sport, work or school decisions, which can be highly problematic. An objective technique to detect the impact of mTBI on the brain is needed. MRI-based T2 relaxation is a quantitative imaging technique that is susceptible to detecting fluid properties in the brain and is a promising marker for detecting subtle neuroinflammation. This study aimed to investigate the potential of T2 relaxometry MRI in assessing mTBI at the individual level.

The current study included 20 male participants with acute sports-related mTBI (within 14 days post-injury) and 44 healthy controls. We statistically compared each mTBI participant’s voxel-wise T2 relaxometry map with the average of controls using a voxel-wise z-test with false discovery rate correction. In addition, five participants were re-scanned after clinical recovery, and their acute scans were compared to their recovery scans.

Results revealed significantly increased T2 relaxation times in 19/20 (95%) of mTBI individuals, compared to controls, in multiple regions, including the hippocampus, frontal cortex, parietal cortex, insula, cingulate cortex and cerebellum. This suggests the presence of increased cerebral fluid in individuals with mTBI. Longitudinal results indicated a partial reduction in T2 relaxation for all five participants, suggesting a resolution over time.

This research highlights the potential of T2 relaxometry MRI as a non-invasive method for assessing neuroinflammation in mTBI. Identifying and monitoring neuroinflammation could aid in predicting recovery and developing individualised treatment plans for individuals with mTBI. Future research would benefit from repeating all MRI scans at recovery to evaluate whether T2-relaxometry normalises or persists.

## Introduction

Approximately 69 million people worldwide experience a mild traumatic brain injury (mTBI) each year.^4^ Whilst in many cases, people recover well, others can experience chronic, disabling impacts that affect their daily functioning.^1,2^ mTBI includes injuries such as concussions and is the most common form of TBI across all ages, with between 70% and 90% of all TBI being considered mild. ^2–4^ mTBI occurs when a person experiences a strong force on the head that causes the brain to move within the skull.^2^ The predominant mechanism of injury is a sudden impact, rotational force or rapid deceleration or acceleration of the brain.^2,5^ The primary injury in mTBI is from the immediate mechanical damage to the brain, resulting in focal and diffuse damage that includes lacerations and contusions and can damage blood vessels, axons and nerve cells.^5^ Following this primary injury, a cascade of secondary injuries can occur. These include inflammation, excitotoxicity, metabolic disturbances, vascular damage, increased intracranial pressure, and blood-brain barrier disruption.^5–8^

The term ‘mild TBI’ may not correspond to the personal experience of many people, as up to 50% of those with an mTBI experience long-lasting symptoms that persist beyond the two to four weeks ‘standard’ recovery time.^9^ The symptoms that accompany the secondary injuries can, if left untreated, last for weeks, months or even years.^7^ Common symptoms of mTBI include headaches, difficulty concentrating, sleep difficulties, a foggy feeling, vestibular disorders, confusion, slowed reaction times, slurred speech, nausea, changes in vision, sensitivity to light, loss of smell or taste and irritability.^2,3,10^ These symptoms can impact patients’ lives in diverse ways. In the longer term, mTBI has been associated with reduced work productivity and increased risk of psychiatric and neurodegenerative diseases.^11,12^

Neuroinflammation is one of the primary drivers behind the secondary injuries associated with brain injuries.^3,13^ The initial tissue damage triggers the activation and recruitment of immune cells by facilitating the production of cytokines and chemokines.^6^ The effect of this inflammatory response is to limit the spread of injury in the brain and to restore homeostasis.^5^ This response has evolved as a protective mechanism, with glial cells working to recover neuronal function in the brain after injury. Microglia play a key role in responding to inflammatory events as they survey the brain, look for structural abnormalities, and work to isolate damaged regions of the brain to prevent the spread of injury. Shortly after the injury, microglia release proinflammatory cytokines, a vital and adaptive part of neuronal preservation.^5^ Neuroinflammation can be beneficial in the acute phase following brain injury, promoting repair of the damaged tissue, possibly inducing neurogenesis and reducing the risk of infection.^14^ However, when this inflammation occurs in excess, it can contribute to the loss of neurons and death of brain tissue that would otherwise be salvageable, resulting in a somewhat preventable secondary injury cascade.^3,6,15–17^

T2 relaxometry is a novel MRI method that can assess microstructural tissue alterations, such as edema, that can accompany different neurological diseases and conditions. This type of MRI provides a marker of intracellular and extracellular injury^18^, such as in epilepsy.^19,20^ The most important property to consider that impacts T2 relaxation time is the water content of the tissue. This water can consist of free water molecules, which are smaller and have a faster spin frequency and a longer T2 relaxation time. Or, the water content can be bound with larger macromolecules with a spin frequency comparable to the Larmour frequency with shorter T2 relaxation times. In regular, healthy tissue, these two types of water exist in equilibrium, but in many pathological conditions (e.g. inflammation), the bound water is released and the free water increases. This creates an inefficient medium for T2 relaxation; therefore, T2 relaxation time increases as tissue water content increases and can indicate pathology.^21–23^

In mTBI patients, brain regions with higher T2 relaxometry signals could reflect neuroinflammation caused by the secondary damage to the brain. Previous research has applied quantitative T2 MRI in other brain disorders, such as epilepsy; however, few studies have used this technique with mTBI. Pedersen et al.^24^ identified brain inflammation in a professional Australian Rules football player who had incurred multiple mTBIs, demonstrating the potential promise of this technique. This study found that abnormally elevated T2 relaxometry persisted throughout each MRI scan while the patient was symptomatic. T2 relaxometry recovered towards the patient’s baseline between the mTBIs but was still elevated compared to the baseline on the last ‘recovery’ scan. Furthermore, T2 relaxometry is a marker of TBI in animal studies. For example, Yang et al.^25^ used a mouse model of mTBI to show that T2 relaxometry can be a valuable tool for identifying acute neurostructural perturbations post-mTBI.

Standard clinical neuroimaging post-mTBI is often negative, with no clinically significant findings.^26^ This can perpetuate the notion that the injury did not result in neuronal pathology despite the patient presenting as symptomatic. Ultimately, there is a need for objective markers of injury so that diagnosis and treatment do not rely exclusively on subjective self-report, which can be susceptible to inaccuracies or reporting bias. There is a need for an advanced method of quantitative MRI that can detect subtle changes in the brain acutely after mTBI. Advanced T2 MRI methods may uncover individual abnormalities indicative of neuroinflammation despite clinically normal radiology findings. In this context, T2 relaxometry could indicate areas of transient neuroinflammation post-mTBI and validate subjective symptom reports or indicate pathology despite a lack of subjective symptoms. Subsequently, acute T2 relaxometry measures could be utilised to detect injury and predict recovery outcomes from mTBI based on the quantified fluid and suspected inflammation in the brain upon acute presentation.

The current study comprises a series of 20 acute mTBI cases individually compared to a control group. This technique will enable a detailed look at potential brain abnormalities on an individual level, enabling any findings to be considered in the context of the patient’s clinical presentation. Deepening our understanding of the underlying pathology of mTBI could contribute to individualised treatment plans and improved patient recovery outcomes.

## Materials and Methods

### Participants

This study consists of 20 male sports players (*M* = 21.6 years old [16-32], *SD* = 4.76) with acute (≤ 14 days) sports-related mTBI. Thirteen mTBI participants were rugby union players, two were football players, one was a hockey player, one was a futsal player, one was a gymnast, one was a surfer and one did jiu-jitsu. We also included 44 male controls that had not suffered an mTBI in the last 12 months and had no lingering symptoms from an mTBI (*M* = 23.3 years old [16-30], *SD* = 4.38). The age of individual mTBI participants and the average age of controls may vary slightly between the 20 individual statistical tests, as our hypothesis is to conduct individual-specific analyses rather than estimate between-group differences. We do not disclose participants’ specific age to minimise the risk of re-identification of participants. Fifteen mTBI participants were recruited at the Axis Sports Concussion Clinics in Auckland, New Zealand and five participants were recruited via community links (e.g. physiotherapists, word of mouth, digital and print advertisements). The control group were recruited through print advertisements at the Auckland University of Technology and The University of Auckland, as well as social media advertisements. Ethics approval was obtained from the Health and Disability Ethics Committee (HDEC – 2022 EXP 11078), New Zealand.

### Magnetic Resonance Imaging Procedure

All magnetic resonance images were acquired using a 3T Siemens MAGNETOM Vida fit scanner (Siemens Healthcare, Erlangen, Germany) located at the Centre for Advanced Magnetic Resonance Imaging (CAMRI) at The University of Auckland, New Zealand, using a 20-channel head coil. A T2 mapping sequence was collected to investigate the anatomical T2 relaxometry. The T2 maps were acquired using an 8 echo Carr-Purcell-Meiboom-Gill (CPMG) sequence (TEs = 28.9, 57.8, 86.7, 115.6, 144.5, 173.4, 202.3 and 231.2 ms; TR = 6s; slice thickness = 2.0mm; voxel size = 2.0 x 2.0 x 2.0mm; matrix size = 112 x 128 x 63; flip angle (FA) = 180°; base resolution = 128; phase resolution = 100%; phase field of view (FOV) = 87.5%). Total T2 mapping acquisition time was 12:02 min.

T1-weighted anatomical images were collected for quality control and normalisation purposes. The T1 weighted images were acquired using a magnetisation-prepared rapid gradient echo (MPRAGE) sequence (TR = 1.9s; TE = 2.5ms; TI = 979ms; FA = 9°; slice thickness = 0.9mm; voxel size = 0.4 x 0.4 x 0.9 mm; matrix size = 192 x 512 x 512; phase FOV = 100%). Total T1-weighted acquisition time was 4:31 min.

A radiologist reviewed clinically relevant MRI images from each participant to check for clinically significant abnormalities that might require further attention. In addition to the acute MRI scans conducted within 14 days post-mTBI, we re-scanned five mTBI participants after they had reached clinical recovery. Follow-up Brain Injury Screening Tool (BIST) questionnaires were completed to determine recovery.

### Data Processing and Statistical Analysis

All MRI images were received in *DICOM* format, converted to *NIfTI* format, and arranged according to the *Brain Imaging Data Structure (BIDS).* ^27^ Image quality assurance checking was conducted in the *MR View* toolbox of *MRtrix* 3^28^ by two investigators in the study (MJB, MP) using the participants’ T1 weighted image as the underlay and their T2 map as the overlay to check for artifacts or abnormalities caused by scanning or processing. Then, preprocessing was conducted on the T2 maps using *PyCharm* version 2022.2.3. This preprocessing step included brain extraction (skull stripping) using the *bet* function in FSL^29^ before normalising each image to the MNI standard space by registering them to a MNI152 2.0 x 2.0 x 2.0mm template image using *FSL FLIRT*.^30^ For each subject, the third T2 echo volume (86.7 ms) was extracted to generate a group average brain image and binary grey matter, white matter, and cerebrospinal fluid masks. Lastly, a monoexponential function with a baseline at each voxel was manually fitted across all eight echo-times using *qMRLAb* in *Matlab R2022B* ^31^ to calculate the T2 relaxation time for each participant^24^.

We conducted a z-test to quantify differences between individual mTBI subjects and controls. Before statistical analysis, spatial smoothing for each image was conducted using a 6mm full-width-half-maximum (FWHM) kernel. All voxels residing within the grey matter mask were considered for final analysis. Given that the main statistical assumption of a z-test is that the underlying data distribution is Gaussian with a mean of 0 and standard deviation of 1, we used a Rank-Based Inverse Normal Transformation^32^ to normalise the data. Here, all T2-relaxometry voxels for each image were ranked and transformed into a Gaussian shape with a distribution mean of 0 and a standard deviation of 1. Next, we obtained z-score maps by subtracting voxel values between individual mTBI maps and the mean of controls divided by the standard deviation of the controls. This is a similar individual-based statistical approach to previous studies.^24,33,34^ We conducted a one-tailed z-test, interpreting positive values only, as the biology of increased T2 relaxometry is well understood (i.e., increased water properties in tissue^18,21–23^). In contrast, the interpretation of decreased T2 relaxometry is less well understood. Additionally, we used False Discovery Rate (FDR)^35^ to correct for multiple comparisons in the z-score maps using a threshold of *q* <0.05. This method obtained z-values and relaxometry times (in ms) for each FDR-based significant cluster, individually for each mTBI participant. Lastly, to determine if the significant T2 relaxometry clusters had resolved with clinical recovery, we subtracted the voxel-wise z-maps of the five participants with recovery MRI re-scans from the voxel-wise z-maps of their acute MRI scans. See Fig. 1 for a schematic overview of our processing and analysis pipeline.

**Figure 1.**
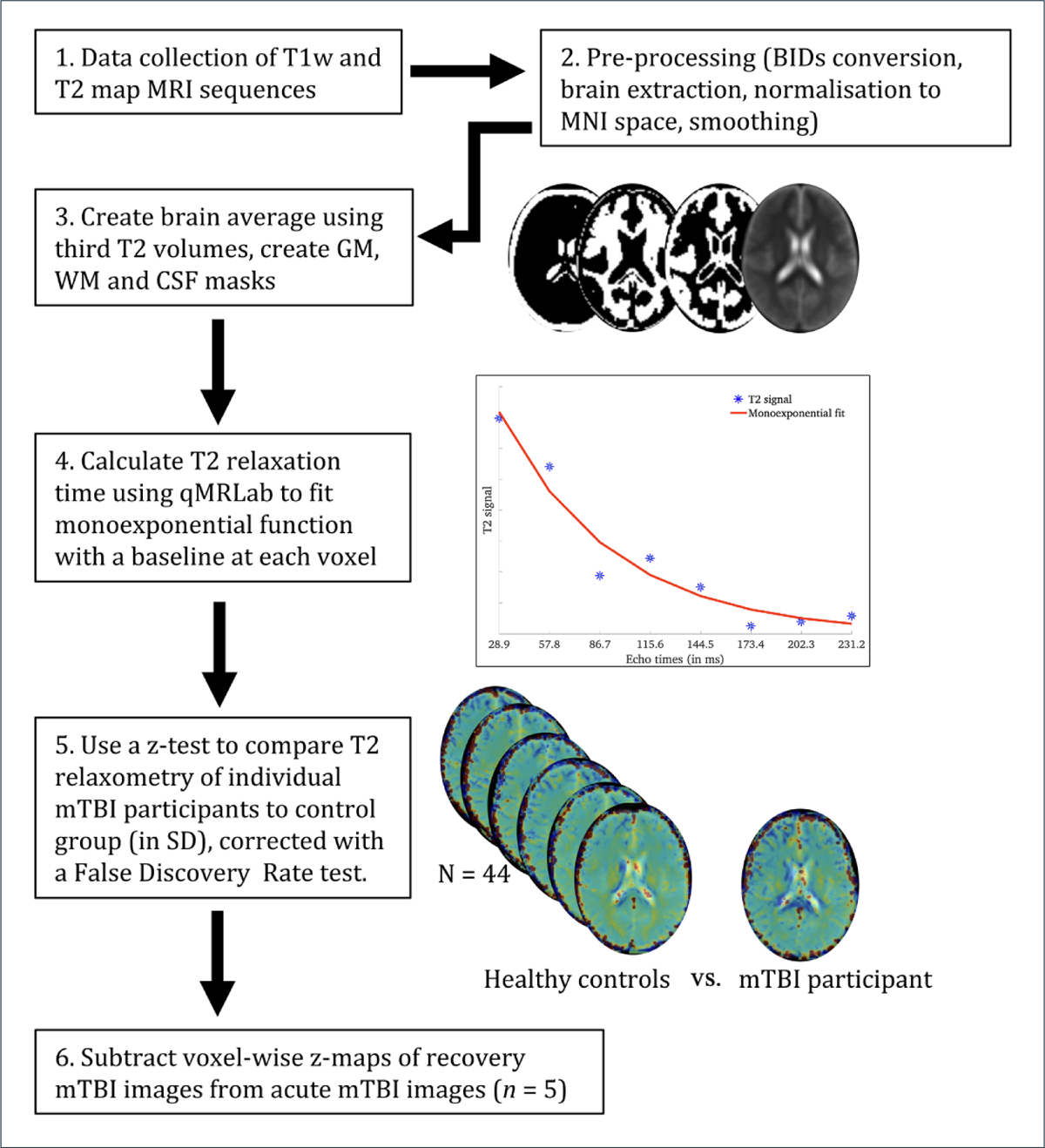
Methodology for MRI Data Processing and Analysis

## Results

### Summary of Acute Scan Findings

The quantitative T2 analysis indicated regions with significantly higher T2 relaxometry times in 19/20 mTBI participants (95%), detailed in Table 1. A radiologist analysed each participant’s MRI scans, and no clinically significant findings were reported. While several significant voxel clusters, such as the cingulate cortex, superior parietal cortex and insula, were shared across multiple participants (see Fig. 2), most of the findings were individual to each case study. These results suggest possible inflammation in the brains of almost all the mTBI participants acutely following their injury. Recovery re-scans indicated either a full reduction (i.e. in all significant acute regions) or a partial reduction (i.e. for some areas but not others) in T2 relaxometry for all five re-scanned participants. The remainder of the Results section will provide comprehensive details about the five case studies that had recovery re-scans, with information regarding their injury, symptoms they presented with, and specific T2 relaxometry results for some areas of their brains. For an extensive description of the remaining 15 participants, see the Supplementary Materials.

**Figure 2.**
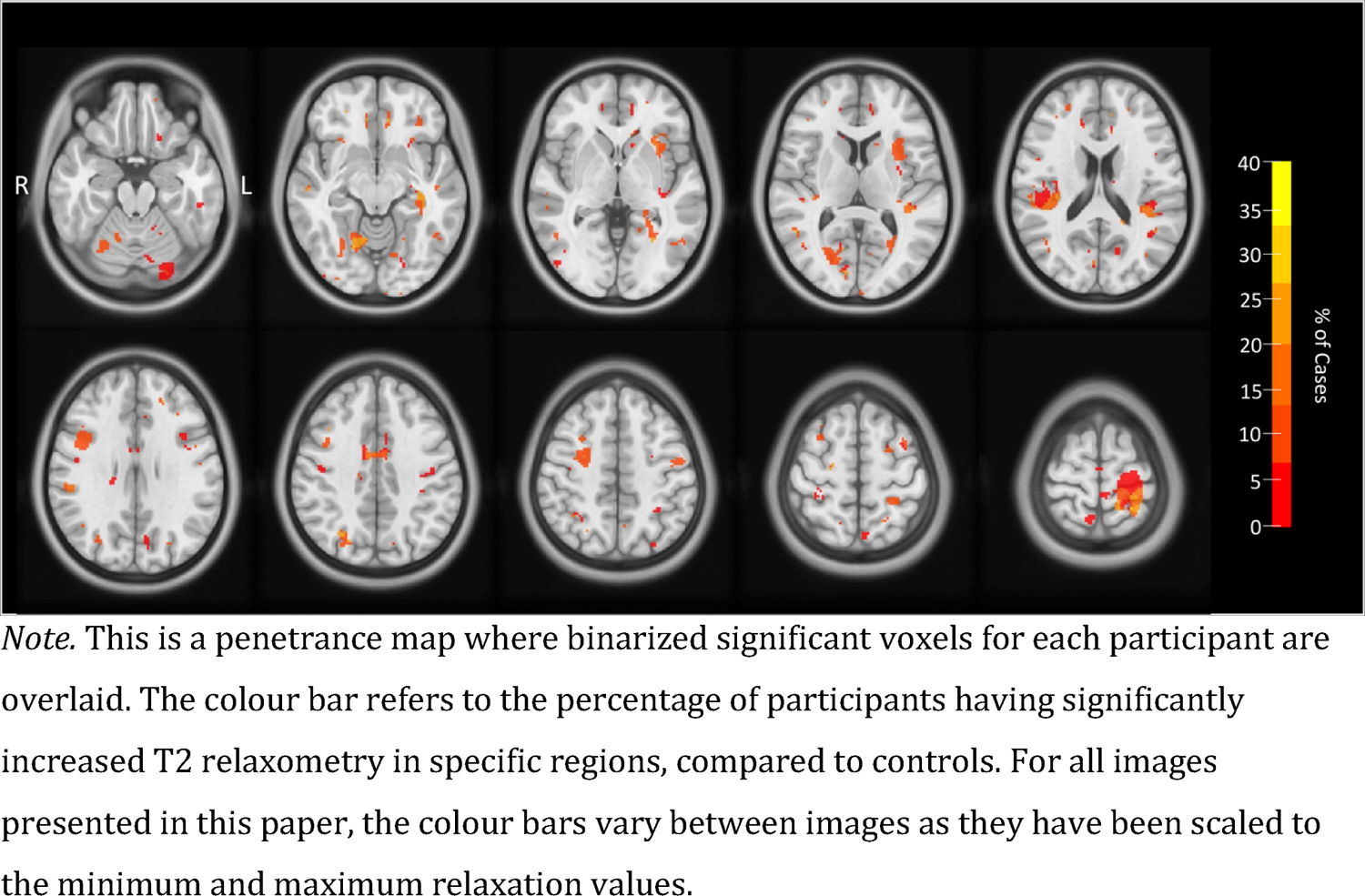
Common Regions of Significant T2 Relaxometry

**Table 1.** Individual Results Showing Voxel Clusters in mTBI with Significantly Higher T2 Relaxometry Times Compared to Controls.

| Case | Region(s) | Raw T2 Relaxometry (ms) | Z-score compared to controls (SD) | Subjective Symptom Reports During & Post-injury |
| --- | --- | --- | --- | --- |
| 1 | L superior parietal<br>L lateral superior frontal<br>L medial superior frontal<br>L orbitofrontal cortex | 146.50<br>2.69e+06*<br>74.18<br>79.78 | 8.40<br>4.08<br>4.38<br>6.50 | Headaches, tiredness, emotional, feeling like he was "in a dream" |
| 2 | L superior parietal<br>L anterior insula<br>R superior frontal | 63.67<br>63.72<br>72.10 | 9.39<br>18.65<br>6.91 | Headaches, nausea, fatigue |
| 3 | R somatosensory cortex<br>R intraparietal sulcus | 76.10<br>51.54 | 8.66<br>15.69 | None reported |
| 4 | L anterior insula<br>L posterior cerebellum | 68.26<br>66.95 | 12.12<br>7.04 | Visual disturbances (seeing "stars"), unsteady, headaches, sensitivity to light |
| 5 | R anterior parietal | 55.86 | 12.83 | Impaired balance, disoriented, feeling "not well", visual disturbances |
| 6 | L hippocampus<br>L lateral occipital | 95.22<br>71.83 | 5.00<br>9.33 | Vomited, ringing in ears, dizziness, headaches, neck pain, light sensitivity, difficulty focusing |
| 7 | L superior parietal<br>R intraparietal sulcus<br>R temporoparietal junction<br>R superior cerebellum | 73.15e+04*<br>72.34<br>65.49<br>89.35 | 7.26<br>14.39<br>3.40<br>8.72 | Impaired balance, dizziness, headaches, sensitivity to light and noise, difficulty concentrating, irritable |
| 8 | Cingulate cortex<br>L inferior supramarginal gyrus | 73.47<br>76.10 | 8.27<br>16.31 | None reported |
| 9 | L superior parietal<br>R posterior insula | 65.66<br>70.28 | 8.81<br>5.31 | Double vision, fogginess, slowed thinking, indecisiveness |
| 10 | L superior parietal<br>R inferior parietal<br>R superior parietal | 54.89<br>65.95<br>70.83 | 15.92<br>14.48<br>8.46 | None reported |
| 11 | L parahippocampal cortex<br>L orbitofrontal cortex<br>R anterior occipital<br>R orbitofrontal cortex | 76.46<br>6.50<br>79.23<br>74.74 | 10.37<br>4.83<br>10.22<br>4.34 | Dizziness, dazed, headaches, neck pain, sensitivity to light, fogginess, mild cognitive symptoms |
| 12 | R superior cerebellum<br>R superior temporal sulcus | 79.46<br>69.54 | 13.15<br>10.23 | Loss of consciousness, "zoning out", headaches/pressure in the head, difficulty concentrating |
| 13 | L anterior temporal | 125.00 | 4.70 | Dazed, pressure in head, neck pain, fogginess, reduced cognitive sharpness |
| 14 | L inferior supramarginal gyrus<br>L anterior occipital<br>L hippocampus<br>R premotor cortex | 70.15<br>57.68<br>274.06<br>87.64 | 25.44<br>13.83<br>8.38<br>19.60 | None reported |
| 15 | R inferior parietal<br>R posterior parietal<br>R superior cerebellum<br>R medial prefrontal cortex | 70.64<br>66.73<br>104.69<br>83.92 | 12.33<br>19.85<br>5.78<br>8.13 | Ataxia, confusion |
| 16 | R sensorimotor cortex | 74.02 | 14.86 | Impaired balance, blurry vision, "pins and needles" in hands, dizziness, headaches |
| 17 | Cingulate cortex<br>L superior frontal<br>L posterior cerebellum | 99.04<br>259.39<br>1.61e+08* | 11.08<br>6.05<br>5.37 | None reported |
| 18 | L superior frontal | 303.65 | 5.28 | None reported |
| 19 | None | N/A | N/A | None reported |
| 20 | L anterior cingulate cortex | 66.32 | 17.51 | None reported |
|  | L medial frontal cortex | 121.10 | 5.68 |  |
|  | L sensorimotor cortex | 79.65 | 4.57 |  |
|  | L precuneus | 38.98 | 5.49 |  |
|  | L temporoparietal junction | 100.25 | 5.60 |  |
|  | R anterior cingulate cortex | 73.53 | 15.48 |  |
|  | R superior parietal | 22.05 | 4.89 |  |

\*As these peak relaxometry values are very high, they could be coming from voxels outside the brain, in the cerebrospinal fluid. However, parts of the clusters appear to be situated within the brain, so they have been included in the analyses.

### Case 1

Case study one suffered an mTBI as a result of a knee to the head during rugby. He experienced headaches, tiredness, abnormal emotional expression and feeling like he was in a dream. These symptoms largely settled at the clinical assessment four days post-injury, with only mild headaches and tiredness being experienced. Brain Injury Screening Tool (BIST) scores were low, with no individual statements obtaining abnormally high scores and an initial total score of 12/160. This patient has experienced one other mTBI in their life, during primary school. Recovery time for the current mTBI is recorded as 20 days. Four significant voxel clusters are apparent when assessing the T2 relaxometry MR images (see Fig. 3). There is a significant cluster in the left superior parietal region, with relaxometry times up to 8.40 SD higher than the controls (peak relaxometry value = 146.5 ms). Two significant clusters are found in the left superior frontal region, the first more laterally with relaxometry times up to 4.08 SD higher than controls (peak relaxometry value = 2.7e+06 ms). As the peak relaxometry value is very high, it could indicate that this peak value is coming from a voxel situated outside the brain, in the cerebrospinal fluid. However, some parts of the cluster appear to be located within the superior frontal region of the brain. The second left superior frontal region is more medial, with relaxometry times up to 4.38 SD higher than controls (peak relaxometry value = 74.2 ms). Lastly, there is a significant cluster in the left orbitofrontal cortex, with relaxometry times up to 6.50 SD higher than controls (relaxometry value = 79.8 ms). After recovery, re-scans indicated that T2 relaxometry was reduced by 14.09 SD in the left superior parietal region, 2.01 SD in the left lateral superior frontal region, and 4.52 SD in the left orbitofrontal cortex. No significant reduction was found for the left medial superior frontal region cluster.

**Figure 3.**
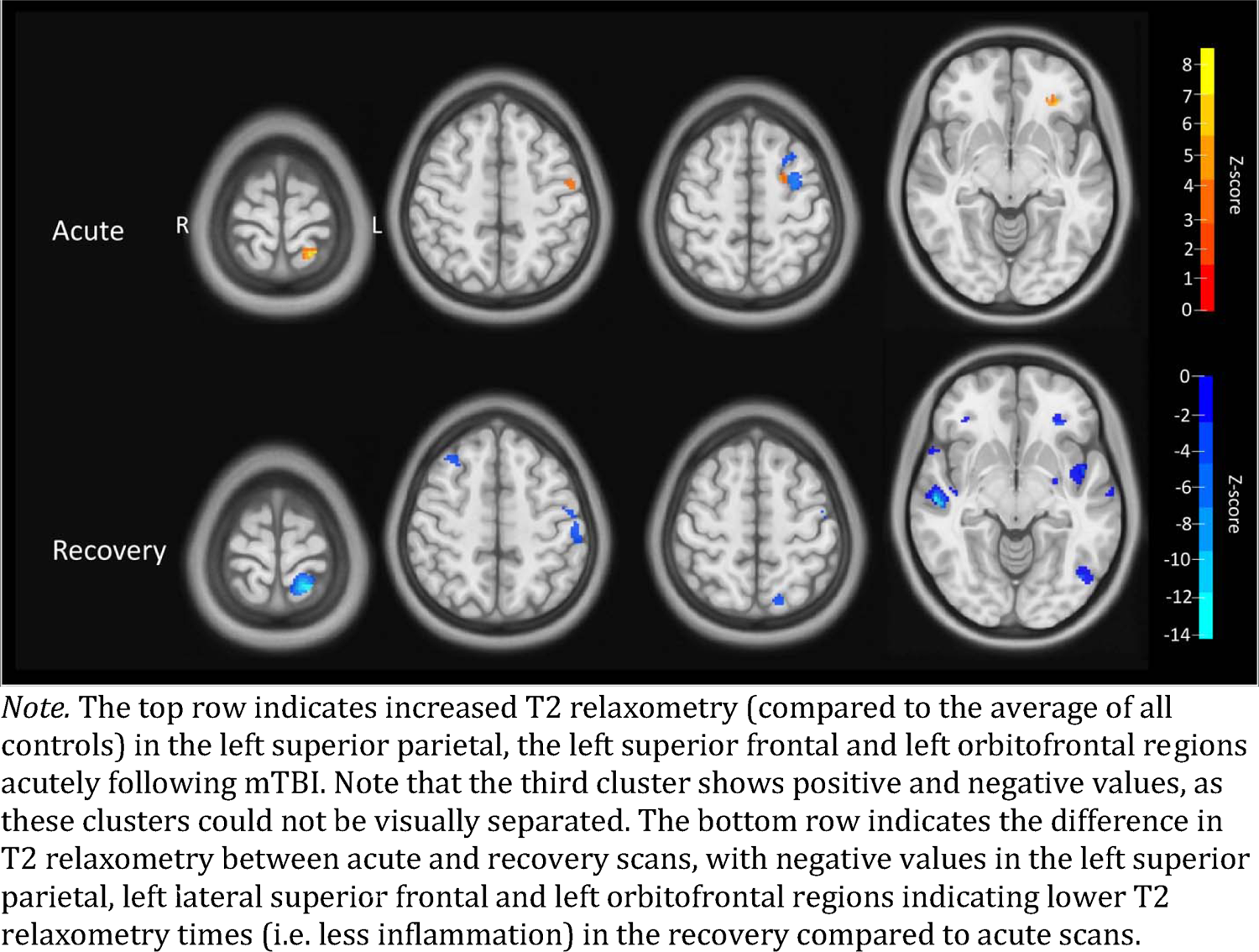
Case 1 T2 Relaxometry Acutely Following mTBI (top) and at Recovery (bottom) (in SD)

### Case 2

Case study two suffered an mTBI when a ball hit his left eye during a hockey game. He experienced developing headaches and nausea after the match and reported mild headaches and fatigue at his clinical assessment nine days post-injury. The patient’s BIST scores were low, with no symptom domains or individual statements obtaining abnormally high scores and an initial total score of 6/160. Recovery time for this injury is recorded as 31 days. Furthermore, he suffered one previous mTBI in 2022. Three significant voxel clusters were identified when analysing the T2 relaxometry images (see Fig. 4). The first is in the left superior parietal region, with relaxometry times up to 9.39 SD higher than controls (peak relaxometry value = 63.7 ms). There are also significant clusters in the left anterior insula, with relaxometry times up to 18.65 SD higher than controls (peak relaxometry value = 63.7 ms) and in the right superior frontal region, with relaxometry times up to 6.91 SD higher than controls (peak relaxometry value = 72.1 ms). After clinical recovery, re-scans indicated that T2-relaxometry was reduced by 8.66 SD in the left superior parietal region, 40.88 SD in the left anterior insula, and 6.15 SD in the right superior frontal region.

**Figure 4.**
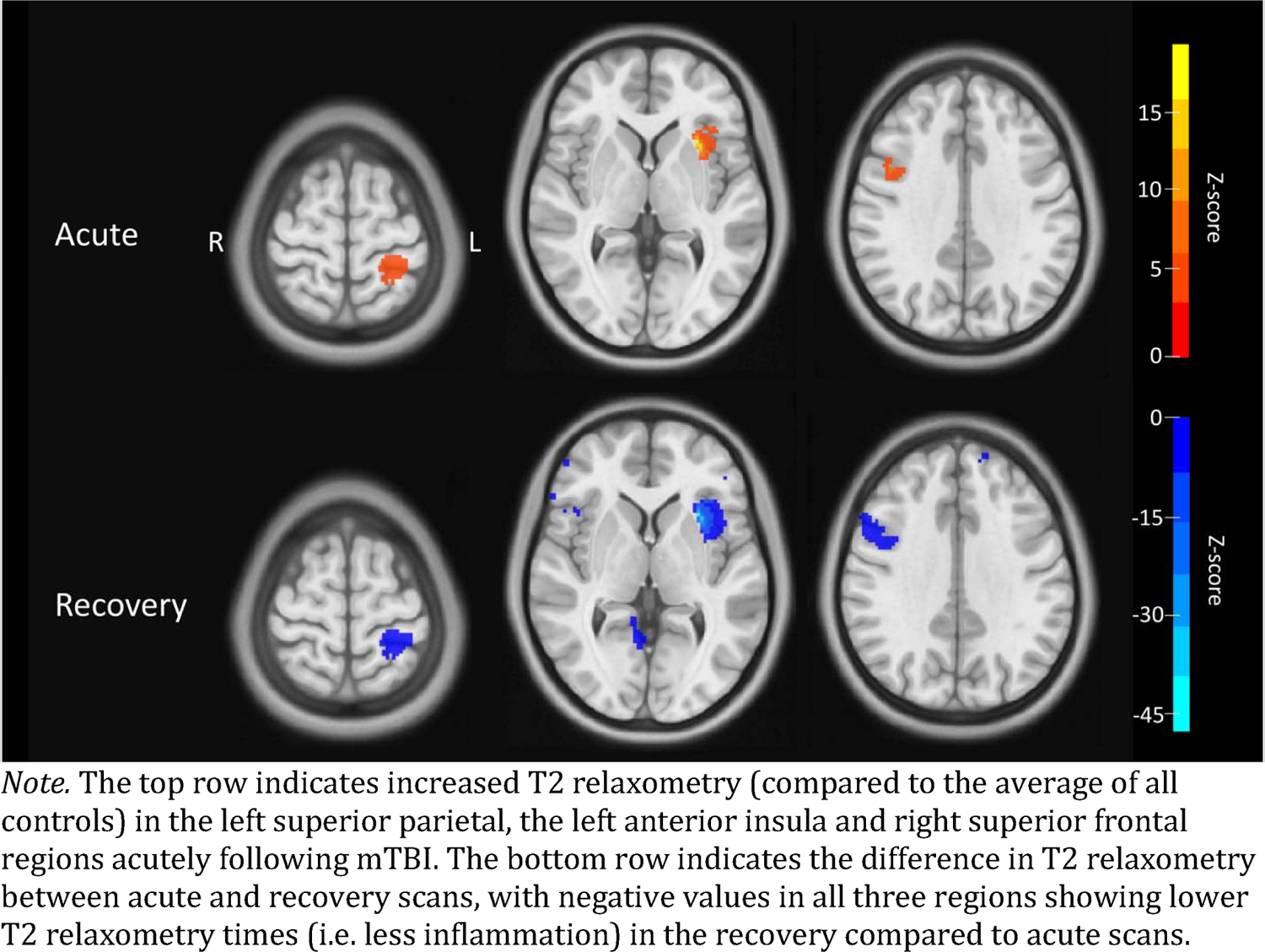
Case 2 T2 Relaxometry Acutely Following mTBI (top) and at Recovery (bottom) (in SD)

### Case 3

Case study three suffered an mTBI when he collided with another player during a football game. The patient did not report any specific symptoms during his clinical assessment 11 days post-injury. The patient’s BIST scores were low with no symptom domains or individual statements obtaining abnormally high scores and an initial total score of 13/160. The patient reported no previous mTBIs and the recovery time for the current mTBI was recorded as 27 days. Two significant voxel clusters are identified when analysing the T2 relaxometry images (see Fig. 5). The first is in the right somatosensory cortex in the parietal lobe, with relaxometry times up to 8.66 SD above controls (peak relaxometry value = 76.1 ms) and the second is in the right intraparietal sulcus, with relaxometry times up to 15.69 SD higher than controls (peak relaxometry value = 51.5 ms). After clinical recovery, re-scans indicated that T2-relaxometry was reduced by 1.54 SD in the right somatosensory cortex and by 0.06 SD in the right intraparietal sulcus.

**Figure 5.**
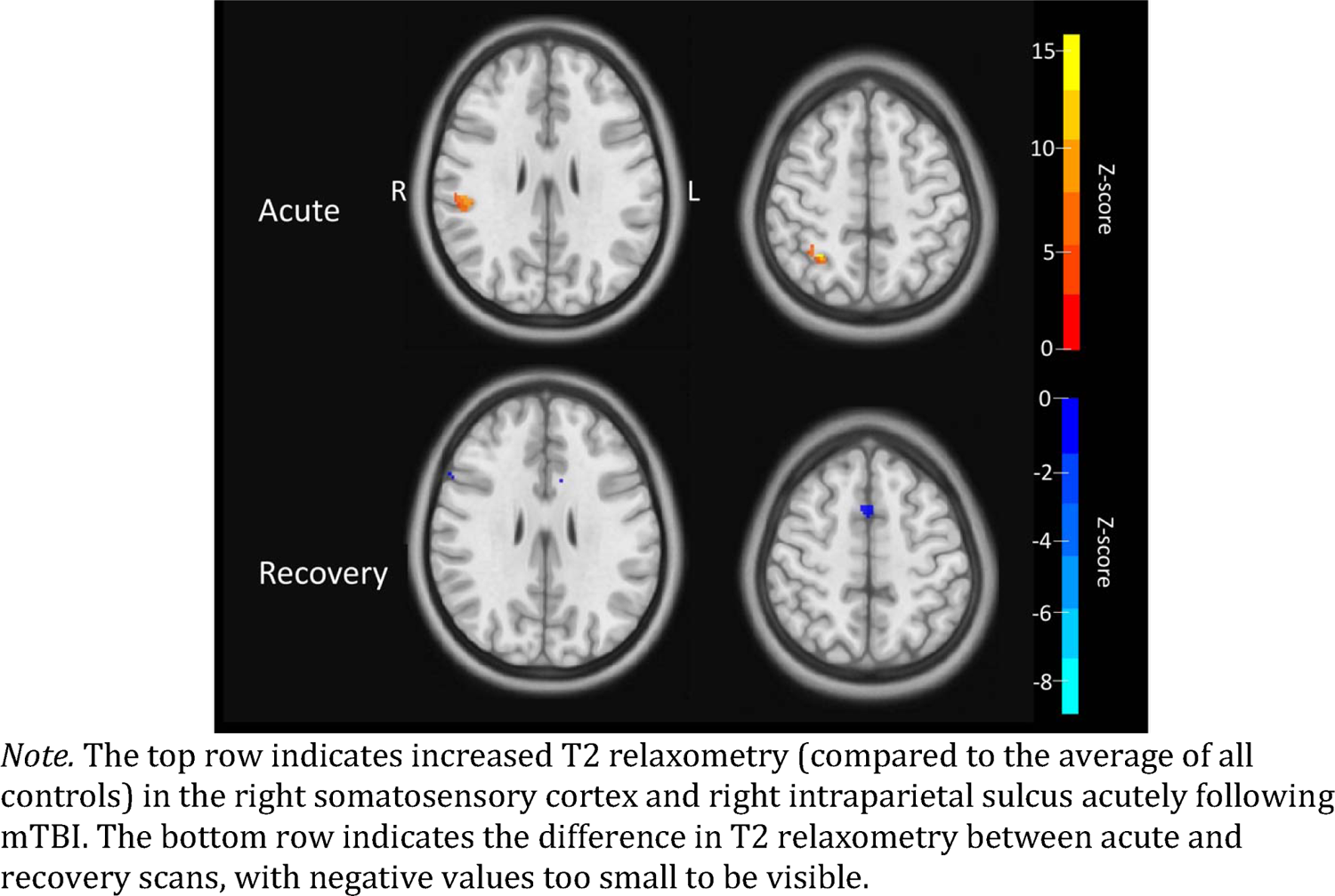
Case 3 T2 Relaxometry Acutely Following mTBI (top) and at Recovery (bottom) (in SD)

### Case 4

Case study four had a head-to-head collision with a teammate during a rugby game and suffered an mTBI as a result. At the time of injury, the patient reports “seeing stars” and being unsteady, followed by headaches and sensitivity to light. Symptoms resolved within a few days and were exacerbated approximately nine days post-injury during Jujitsu training. At his clinical assessment, 12 days post-injury, the patient reports headaches and fatigue. BIST scores were low, with no symptom domains or individual statements obtaining abnormally high scores and an initial total score of 22/160. The patient has reported one previous mTBI, in 2021. Recovery time for the current mTBI is recorded as 26 days. When analysing T2 relaxometry MRI data for this patient, two significant clusters of voxels were discovered (see Fig. 6). The first is in the left anterior insula, with relaxometry times up to 12.12 SD higher than controls (peak relaxometry value = 68.3 ms) and the second is in the left posterior cerebellum, with relaxometry times up to 7.04 SD above controls (peak relaxometry value = 67.0 ms). After clinical recovery, re-scans indicated that T2-relaxometry was reduced by 11.22 SD in the left anterior insula, but no reduction was found in the left posterior cerebellum.

**Figure 6.**
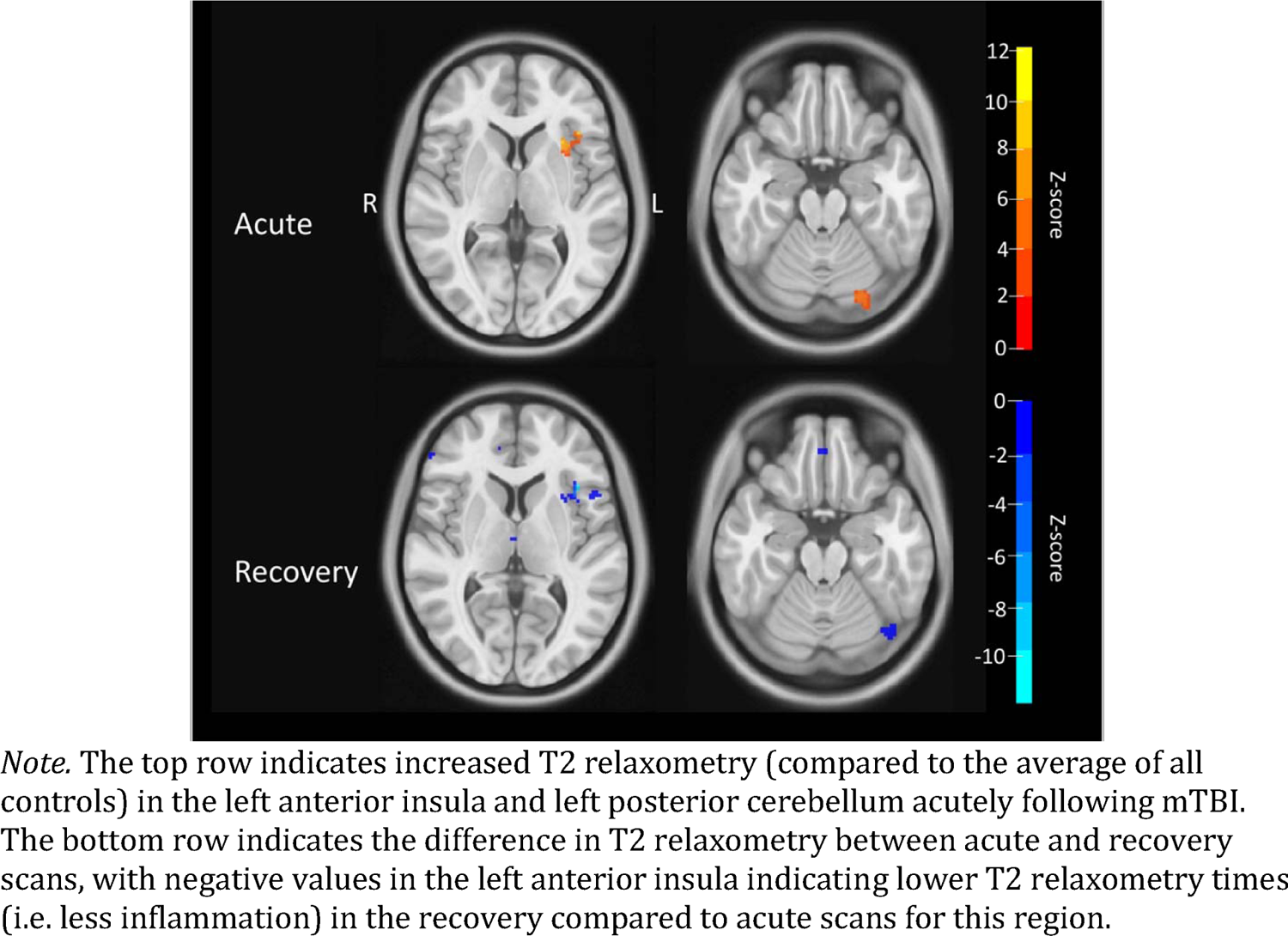
Case 4 T2 Relaxometry Acutely Following mTBI (top) and at Recovery (bottom) (in SD)

### Case 5

Case study five sustained an mTBI during gymnastics training when he fell off a high bar. The patient reports stumbling when returning to his feet but not experiencing any other symptoms at the time of injury. The following day, he reports feeling disoriented and unwell. Two days after the first injury, the patient again falls on his head during gymnastics training. This time, the patient reports seeing flashes of colour and blurred vision. No additional symptoms are reported during his clinical assessment seven days post-injury. BIST scores were low with no symptom domains or individual statements obtaining abnormally high scores and an initial total score of 34/160. Recovery time for this injury is recorded as 56 days and this patient has suffered one previous mTBI, in approximately 2017/2018. When analysing the T2 relaxometry images, one significant cluster of voxels was found in the right anterior parietal lobe (see Fig. 7), with relaxometry times of up to 12.83 SD above controls (peak relaxometry value = 55.9 ms). After clinical recovery, re-scans indicated that T2-relaxometry was reduced by 12.68 SD in this region.

**Figure 7.**
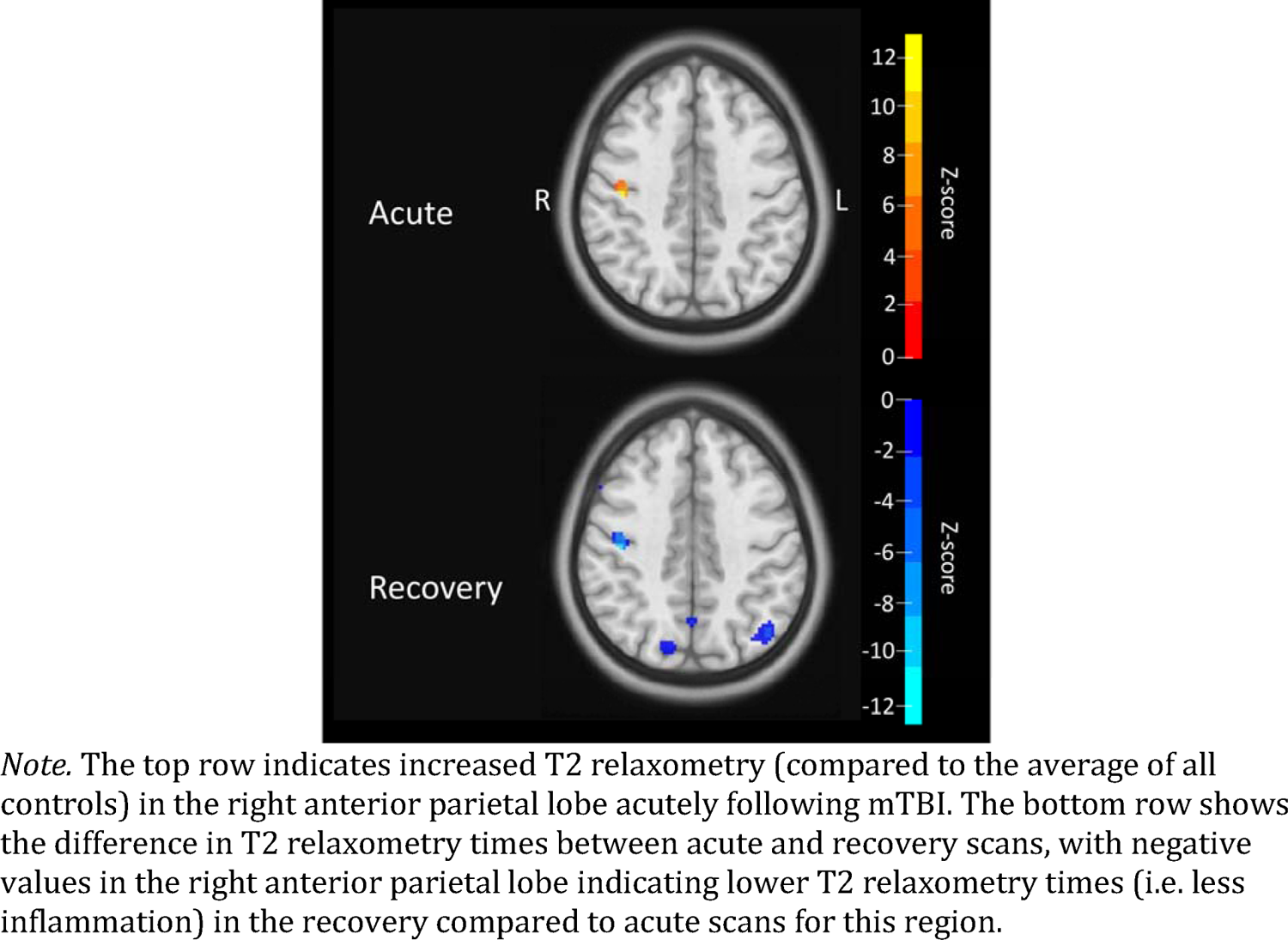
*Case 5 T2 Relaxometry Acutely Following mTBI (top) and at Recovery (bottom) (in SD)*

## Discussion

### Individual-level Brain Abnormalities in Acute mTBI

Group-level statistical analyses dominate clinical neuroscience, and while they have utility in enabling reliable comparisons between groups, they may not detect brain abnormalities unique to individuals. Our study compared each mTBI patient to the average of controls, enabling a comprehensive analysis of significant voxel clusters for specific individuals. We found that 19/20 mTBI patients had significantly higher T2 relaxometry times than the control group (see Fig. 3-7 for examples). Regions with significantly higher T2 relaxometry are summarised in Table 1 and include the left hippocampus, left superior parietal cortex, left superior frontal cortex, left orbitofrontal cortex, left supramarginal gyrus, left anterior insula, left posterior cerebellum, right temporoparietal junction, right superior cerebellum, right medial prefrontal cortex, right somatosensory cortex, and the cingulate cortex. Given T2 relaxometry’s sensitivity to quantify water properties, we believe our findings indicate signs of subtle neuroinflammation during acute stages of mTBI.

An advantage of investigating brain abnormalities at an individual level is the potential to draw possible links between objective MRI findings and clinical symptomatology. We observed that there is an overlap between mTBI symptoms and T2-relaxometry findings. For example, the abnormal emotional expression case one experienced directly following his injury may be related to possible inflammation in the orbitofrontal cortex (see Fig. 3). It is also possible that the “unsteady” feeling that case four experienced post-mTBI is linked to inflammation in the cerebellum (see Fig. 6). In addition, it is possible that case six’s issues with memory and vision, as indicated by the BIST, have neural bases involving elevated T2-relaxometry in hippocampal and occipital regions (see Supplementary Materials). Lastly, the dizziness and impaired balance experienced by case seven could be related to possible inflammation in the cerebellum (see Supplementary Materials). Ultimately, further investigation would be required to ascertain whether these potential regions of neuroinflammation were responsible for the subjectively reported symptoms.

A radiologist reviewed MRI scans from all participants, and none were deemed to have any findings that would warrant clinical follow-up. This indicates that the patients in this study have potentially pathological brain abnormalities despite no findings on hospital-based MRI protocols. Of note, the radiologist’s report for case 18 did indicate a structural brain change in the left superior frontal gyrus (see Supplementary Materials). This region was not deemed clinically significant by the radiologist and likely unrelated to the current brain injury. Still, this region spatially aligned with our findings of a significant T2 relaxometry voxel cluster in the left superior frontal lobe and further supports our hypothesis that T2 relaxometry can identify structural brain abnormalities. This further demonstrates the utility of T2 relaxometry for uncovering subtle abnormalities.

### Common Regions of Increased T2 Relaxometry across mTBI Individuals

As well as individual variation in T2 relaxometry times, we observed a range of significant voxel clusters that multiple participants had in common, affecting up to 40% of mTBI cases (Fig. 2). These regions include the cingulate cortex, anterior insula, cerebellum and superior parietal cortex. Regions with increased T2 relaxometry times across multiple participants could indicate that specific brain regions are more susceptible to acute brain changes following mTBI. If there were regions that appear more vulnerable, these regions could serve as potential biomarkers for injury and predictive indicators for recovery from mTBI for future players. For example, inflammation in specific brain regions acutely following injury may be linked to poorer recovery. Therefore, early identification of this inflammation could facilitate individualising treatment plans (e.g., a more conservative return to sport) and improve recovery outcomes.

It is also worth considering whether specific brain regions are selectively vulnerable after a brain injury. Previous meta-analytic research suggests that insular and cingulate regions are preferentially vulnerable to grey matter loss in a range of neuropsychiatric conditions,^36^ being regions containing specialised long spindle-shaped neurons known to be susceptible to neurodegenerative disease.^37^ The insular cortex also appears to be linked to the inflammatory response, which could explain why it is one of the most common regions that show increased T2 relaxometry times in participants. Rolls^38^ explains that the role of the insular cortex in immune regulation is becoming clear, as this region seems to play a role in forming the brain’s representation of the immune state. This may be partly due to the insular cortex’s anatomical connections to the peripheral sensory and autonomic nervous systems.^38^ Furthermore, Mansson et al.^39^ explain that anterior insula morphology might be a factor in the sensitivity to develop symptoms of sickness and anxiety in response to inflammation. In particular, a larger anterior insula was linked to having a stronger response to sickness.^39^ Additionally, the insular and cingulate cortex may be functionally connected by their role in the inflammatory response^39^, which could be why they are both common regions among participants. Lastly, Koren et al.^40^ also emphasise this connection by demonstrating the importance of the insular cortex for storing information about the inflammatory response. Future research could further investigate possible functional explanations for these common regions to deepen our understanding of the pathology underlying mTBIs.

### Recovered mTBI Participants show Reduced T2 relaxometry

For all five participants with longitudinal data available, MRI recovery re-scans suggest that increased T2 relaxometry present in the acute scans is reduced once they are clinically recovered (see Figs. 3-7). These recovery re-scans provide valuable evidence to support the idea that the potential neuroinflammation found on acute scans was linked to the mTBI, as it seems to (at least partially) resolve as the participant recovers. This evidence aligns with a previous case report suggesting that brain-specific T2 relaxometry reduces after clinical recovery ^24^. Therefore, our findings support the hypothesis that inflammation may occur in the brain acutely following mTBI as part of the overall inflammatory response to the injury. However, this response may be transient. This finding also raises the question of whether mTBI is an acute or chronic injury with regard to brain pathology. While clinical recovery is generally determined by subjective symptom reporting (via symptom questionnaires and/or reports made to doctors), it is unknown whether any objective biomarkers of mTBI can be used to indicate recovery. Conducting recovery re-scans with a larger sample size would enable conclusions regarding whether this pattern is generalisable to the mTBI population. If our findings are replicated at a larger scale, they could be utilised as an objective measure of recovery from mTBI.

### Clinical Implications of Potential Human Neuroinflammation Markers

Conventional MRI images are generally qualitative. Without a quantitative metric to interpret the signal intensities (independent of scanner hardware and sequences), comparing MRI images longitudinally or across subjects is challenging. While the conventional method provides good tissue contrast, the signal intensity can only be interpreted qualitatively. Consequently, interpreting the resulting images relies on selecting an appropriate sequence and a sound understanding of the signal contrast relative to the pathophysiology. By not depending on this subjective process and to obtain quantitative information, the contributions of different contrast mechanisms need to be extracted.^21^ Quantitative measures can isolate the contributions of individual MR contrast mechanisms (i.e. T1, T2 and T2*). The relaxation time is sensitive to water content, iron levels and tissue structure. It measures the biophysical parameters by decoupling the contrast mechanisms contributing to the signal. This makes this method an unbiased metric for comparisons between MRI data.^21^

Using T2 relaxometry MRI analysis enables detailed maps of potential brain inflammation. This method may provide insights into the patient’s injury that other imaging methods would not obtain. Uncovering regions with possible acute inflammation following mTBI could affect the individual’s recovery and return to play. For example, gaining a deeper understanding of brain abnormalities post-injury could help clinicians individualise treatment plans instead of basing the plan on group averages, self-reports, or generic guidelines for recovery. To base treatment decisions on personal data means tailoring the plan for what would benefit the individual - for example, increasing or decreasing the recommended stand-down period before returning to play. Therefore, the chance of being too lenient in the approach to rest and recovery is reduced. Furthermore, knowing whether someone’s brain was showing signs of inflammation could instigate a need for altering the recommended physical activity (as this has been linked to inflammation^16,17,41,42^), or it could entail incorporating other methods for reducing inflammation (e.g. anti-inflammatory medications) that would not have otherwise been suggested. Overall, understanding brain pathology on an individual level and subsequently customising treatment plans could result in a more efficient and safer return to play for athletes with an mTBI.

### Limitations and Future Directions

The current study has several limitations to consider when discussing the results. Firstly, while all MRI scans were conducted during a 14-day window in the ‘acute’ stage following mTBI, there was inter-participant variability in the exact timing and injury severity. Minor timing and severity differences could be a potential explanatory variable to consider for differences in neuroinflammation. Furthermore, the current study only included male participants and the main cohort in this study were rugby players, a male dominant sport. If both sexes were to be included, separating them into different groups and comparing them would be sensible. This would require a larger sample size and reduce the feasibility of the study. Although longitudinal research is challenging (especially as MRI studies are a large commitment for the participants), it would be beneficial for future research to re-scan more mTBI participants after recovery. This way, T2 relaxometry times, and therefore possible sites of neuroinflammation, could be compared in the ‘injured’ and the ‘recovered’ brains of participants to see if significant clusters in the acute scans normalise with recovery.

## Conclusion

We observed that nearly all mTBI participants had evidence of elevated brain T2 relaxometry after an mTBI. We hypothesise that these findings indicate possible neuroinflammation and enable a deeper understanding of their injuries’ pathology. Furthermore, comparing the recovery MRI scans of five participants to their acute scans indicated that T2 relaxometry normalises at recovery. This suggests a possible transient nature to the brain pathophysiology and provides a possible objective imaging marker for reliably judging recovery following mTBI. Quantitative T2 relaxometry methods provide individual, detailed maps of potential brain abnormalities and may offer new insights into a patient’s clinical presentation. Understanding the role that inflammation plays acutely post-mTBI could have implications for individualising treatment approaches (e.g. increasing focus on methods for reducing inflammation) and, therefore, improving recovery outcomes for mTBI patients.

## Data Availability

All data produced in the present study are available upon reasonable request to the authors

Data can be made available by request to the corresponding author.

## Acknowledgements

Thank you to Amabelle Voice-Powell for her contribution to external clinic relationship building and the data collection process, Cassandra Mcgregor for her contribution to the data collection process, and Tania Ka’ai for her contribution to the concept and design of the study. In addition, we would like to thank Axis Sports Concussion Clinic for their assistance with mTBI recruitment and the Centre for Advanced Magnetic Resonance Imaging (CAMRI) for their assistance with MRI data collection.

## Funding

This project was funded by a grant from the Health Research Council of New Zealand (HRC), grant #21/622.

## Competing Interests

None to declare.

## Supplementary Material

### Case 6

Case six suffered an mTBI resulting from a rugby tackle whereby he was dropped on his neck. Clinical reports indicate some vomiting, dizziness and ringing in the ears immediately post-injury, as well as neck pain, headaches, light sensitivity and difficulty focusing four days post-injury. BIST scores are high and indicate a 10/10 rating for the statements “I don’t like bright lights” and “I don’t like loud noises”, as well as a 9/10 rating for the statement “I have trouble with my eyesight (vision)”, an 8/10 score for “I forget things” and a total initial BIST score was 140/160. This is the first mTBI that this patient has experienced, and recovery time was recorded as 26 days. Two significant findings are discovered after examination of T2 relaxometry MR images. Firstly, there is a significant cluster of voxels in the left hippocampal region, with relaxometry times up to 5.00 SD higher than that of the control group (peak relaxometry value = 95.2 ms). Secondly, there is a significant cluster in the left occipital lobe, with relaxometry times up to 9.33 SD higher than controls (relaxometry value = 71.8 ms).

### Case 7

Case study seven received a kick to the head during a rugby tackle and suffered an mTBI as a consequence. Directly after the incident, the patient complained of feeling dizzy and off-balance and at his clinical assessment four days post-injury, he reported headaches, sensitivity to light and noise, as well as difficulty concentrating and some increased irritability. BIST scores indicate a 6/10 rating for the statement “I don’t like bright lights”, a 7/10 for the statement “I don’t like loud noises”, a 6/10 for the statement “It takes me longer to think” and a 7/10 rating for the statement “I have trouble concentrating”. His total initial score was 78/160. The patient has not experienced any previous mTBIs and recovery time is recorded as 47 days. Four significant voxel clusters were discovered when assessing the patient’s T2 relaxometry images. Firstly, there is a significant cluster in the left superior parietal region, with relaxometry times up to 7.26 SD above the control average (peak relaxometry value = 731,456.0 ms). Again, as this peak relaxometry value is very high, it may be coming from a voxel that is situated outside the brain, in the CSF. However, a portion of the significant cluster appears to be situated within the superior parietal region of the brain. Secondly, there is a significant cluster in the right intraparietal sulcus, with relaxometry times up to 14.39 SD higher than controls (peak relaxometry value = 72.3 ms). Next, there is a significant cluster in the right temporal parietal junction, with relaxometry times up to 3.40 SD above controls (peak relaxometry value = 65.5 ms). Lastly, the right superior cerebellum contains a significant cluster with relaxometry times up to 8.72 SD above controls (peak relaxometry value = 89.4 ms).

### Case 8

Case study eight suffered an mTBI during a rugby game, during which he lost consciousness momentarily. No additional clinical data was available for this patient regarding specific symptom complaints during or post-injury. BIST scores indicate a rating of 8/10 for the statement “I don’t like bright lights” and a total initial score of 52/160. This patient has experienced two previous mTBIs, with the last one two months prior in 2023. Recovery time for the current injury is recorded as 27 days. Upon examination of T2 relaxometry images, two significant findings were discovered. The first is in the cingulate cortex, with relaxometry times up to 8.27 SD higher than controls (peak relaxometry value = 73.5 ms) and the second overlaps the left superior Wernicke’s area and inferior supramarginal gyrus, with relaxometry times up to 16.31 SD (peak relaxometry value = 76.1 ms).

### Case 9

Case study nine suffered an mTBI during a rugby game when he fell and hit the back of his head. The patient reported having double vision at the time of injury and then developing brain fog, slowed thinking and indecisiveness three days post-injury. At the time of his clinical assessment, 12 days post-injury, the patient reports no symptoms and a 0/10 rating for all BIST statements. No available data indicates whether the patient has suffered any previous mTBIs. Recovery time for this patient is recorded as 18 days. When looking at the T2 relaxometry images, two significant voxel clusters are found. The first is in the left superior parietal lobe, with relaxometry times of up to 8.81 SD higher than controls (peak relaxometry value = 65.7 ms), and the second is in the right posterior insula, with relaxometry times up to 5.31 SD higher than controls (peak relaxometry value = 70.3 ms).

### Case 10

Case study 10 sustained an mTBI while surfing. This patient reports experiencing a seizure and loss of consciousness with this mTBI. No additional clinical data was available for this patient regarding specific symptom complaints during or post-injury. The patient indicated a 7/10 BIST rating for the statement “I don’t like bright lights”, a 7/10 for “I don’t like loud noises”, a 6/10 for “I feel dizzy or like I could be sick”, an 8/10 for “I feel clumsy”, a 7/10 for “It takes me longer to think”, a 7/10 for “I forget things”, and a 7/10 for “I get confused easily”, with a total initial BIST score of 69/160. The patient reports numerous previous mTBIs, with the exact amount and timing of the most recent one being unknown. The participant did not report the recovery time for the current mTBI. Three significant voxel clusters were located when examining the T2 relaxometry images. The first was a large cluster in the left superior parietal lobe, with relaxometry times up to 15.92 SD higher than controls (peak relaxometry value = 54.9 ms). The second was another large cluster, in the right inferior parietal lobe, with relaxometry times up to 14.48 SD above controls (peak relaxometry value = 66.0 ms) and the last was in the right superior parietal lobe, with relaxometry times up to 8.46 SD above controls (peak relaxometry value = 70.8 ms).

### Case 11

Case study 11 suffered an mTBI in rugby when another player’s shoulder hit the right side of his head. At the time of the injury, the patient complained of feeling dizzy, dazed, with a headache and neck pain. At clinical assessment, 11 days post-injury, he indicated sensitivity to light and feeling “foggy” with mild cognitive symptoms. BIST scores were low with no symptom domains or individual statements obtaining abnormally high scores and an initial total score of 18/160. This patient has suffered four previous mTBIs, with the most recent being in November 2021. The recovery time for the current mTBI is recorded as 24 days. Three significant voxel clusters are apparent when assessing the T2 relaxometry MR images. The first is in the left parahippocampal cortex, with relaxometry times up to 10.37 SD higher than controls (peak relaxometry value = 76.5 ms). Secondly, there is a significant cluster in the left orbitofrontal cortex, with relaxometry times up to 4.83 SD higher than controls (peak relaxometry value = 6.5 ms). Next, there is a cluster in the right anterior occipital lobe, with relaxometry times up to 10.22 SD higher than controls (peak relaxometry value = 79.2 ms). Lastly, there is a significant cluster in the right orbitofrontal cortex, with relaxometry times up to 4.34 SD higher than controls (peak relaxometry value = 74.7 ms).

### Case 12

Case study 12 suffered an mTBI during a rugby tackle, with the left side of his head hitting the ground and the knee or elbow of another player hitting the right side of his head. At the time of injury, he lost consciousness for a few seconds, was reported to be “zoning out” and experiencing headaches. At his clinical assessment, 11 days post-injury, he reports having some difficulty concentrating at school, as well as having headaches and some pressure in his head. BIST scores indicate a rating of 8/10 for the statement “I don’t like bright lights” and a 9/10 for the statement “I don’t like loud noises”, with a total initial BIST score of 61/160. This patient has not experienced any previous mTBIs and recovery time is recorded as 23 days. Two significant voxel clusters are apparent when assessing the T2 relaxometry MR images. These are in the right superior cerebellum, with a relaxometry time up to 13.15 SD higher than the control group average (peak relaxometry value = 79.5 ms) and the right superior temporal sulcus, with relaxometry times up to 10.23 SD higher than controls (peak relaxometry value = 69.5 ms).

### Case 13

Case study 13 sustained two separate head injuries during a rugby game; the first when his head collided with another player’s knee, and then the second when he received an elbow to the left side of his face. The patient reports feeling shocked and dazed at the time of the injuries and reports feeling pressure in his head as well some neck pain, fogginess and disturbance to cognitive functions such as memory four days post-injury. There was no data available for this patient with regards to whether loss of consciousness occurred or whether any previous mTBIs have been suffered. BIST scores were low with no symptom domains or individual statements obtaining abnormally high scores, and an initial total score of 42/160. Recovery time for this patient is recorded as 17 days. One significant voxel cluster was identified when evaluating this patient’s T2 relaxometry MR images. This was in the left anterior temporal region, with relaxometry times up to 4.70 SD above controls (peak relaxometry value = 125.0 ms).

### Case 14

Case study 14 suffered an mTBI during a tackle in a football game when the ball was kicked directly at face, losing vision for a few seconds. No additional clinical data was available for this patient regarding specific symptom complaints during or post-injury. BIST scores indicate a 6/10 rating for the statement “I don’t like bright lights” but low scores for other statements with an initial total score of 61/160. This patient has suffered one previous mTBI in 2018. Recovery time for the current mTBI is recorded as 20 days. Four significant voxel clusters were discovered when analysing the T2 relaxometry MR images. The first area overlaps the left superior Wernicke’s area and inferior supramarginal gyrus, with relaxometry times up to 25.44 SD higher than controls (peak relaxometry value = 70.2 ms). The next overlaps the left posterior corpus callosum and anterior occipital lobe, with relaxometry times up to 13.83 SD higher than controls (peak relaxometry value = 57.7 ms). In addition, there is a significant cluster in the left hippocampus, with relaxometry times up to 8.38 SD higher than controls (peak relaxometry value = 274.1 ms) and in the right premotor cortex, with relaxometry times up to 19.60 SD higher than controls (peak relaxometry value = 87.6 ms).

### Case 15

Case study 15 was hit on the right side of his maxilla by an elbow during a rugby training session, resulting in an mTBI. The patient had ataxia (impaired coordination) and confusion directly following the incident. During his clinical assessment two days post-injury he reports a high symptom load as indicated by vision motion sensitivity testing, for example. However, the patient’s BIST scores were low with no symptom domains or individual statements obtaining abnormally high scores and an initial total score of 56/160. There is no available data for whether the patient has experienced any previous mTBIs and recovery time for this mTBI is recorded as 18 days. Four significant voxel clusters were found when analysing the T2 relaxometry MR images. The first is in the right inferior parietal region, with relaxometry times up to 12.33 SD higher than controls (peak relaxometry value = 70.6 ms). Next, there is a significant cluster in the right posterior parietal region, with relaxometry times up to 19.85 SD higher than controls (peak relaxometry value = 66.7 ms). Furthermore, there is a significant cluster that overlaps the right superior cerebellum and inferior occipital region, with relaxometry times up to 5.78 SD higher than controls (peak relaxometry values = 104.7 ms) and in the right medial prefrontal cortex, with relaxometry times up to 8.13 SD higher than controls (peak relaxometry value = 83.9 ms).

### Case 16

Case study 16 suffered an mTBI during a rugby game when the knee of his opponent made contact with the back of his head. The patient experienced loss of balance at the time of injury and 24-hours-later developed numbness in his hands, blurry vision and headaches, with pins and needles in his hands, dizziness and headaches still at his clinical assessment 11 days post-injury. BIST scores indicate a 7/10 rating for the statement “I feel dizzy, or like I could be sick” and an initial total score of 54/160. Recovery time for this mTBI is recorded as 117 days. Furthermore, the patient has suffered four previous mTBIs, with the latest one occurring in 2021. When analysing the T2 relaxometry images, one significant voxel cluster was identified in right sensorimotor cortex, with relaxometry times up to 14.86 SD above controls (peak relaxometry value = 74.0 ms).

### Case 17

Case study 17 suffered an mTBI during a tackle in a rugby game. No clinical notes are available for this patient in relation to his symptoms at the time of the injury or at the time of clinical presentation. BIST scores indicate a rating of 6/10 for the statement “I don’t like loud noises”, an 8/10 for “It takes me longer to think”, a 9/10 for “I forget things”, a 6/10 for “I get confused easily”, a 6/10 for “I have trouble concentrating”, a 6/10 for “I get angry or irritated easily”, a 7/10 for “I feel tired during the day” and an 8/10 for “I need to sleep a lot more or find it hard to sleep at night”, with an initial total BIST score of 79/160. This patient reports two previous mTBIs, with the most recent mTBI only two months prior in May 2023. Recovery time for the current mTBI is recorded as 66 days. Three significant voxel clusters were identified when analysing the T2 relaxometry MR images. Firstly, in the cingulate cortex with relaxometry times up to 11.08 SD above controls (peak relaxometry value = 99.0 ms), secondly in the left superior frontal lobe with relaxometry times up to 6.05 SD above controls (peak relaxometry value = 259.4 ms) and thirdly in the left posterior cerebellum, with relaxometry times up to 5.37 SD above controls (peak relaxometry value = 1.6e+08 ms).

### Case 18

Case study 18 suffered an mTBI during a rugby game when another player’s knee made contact with his head and he lost consciousness momentarily. No additional clinical data is available for this patient regarding specific symptom complaints during or post-injury.

BIST scores were low with no symptom domains or individual statements obtaining abnormally high scores and an initial total score of 2/160. This patient reports no previous mTBI and the recovery time for the current injury is recorded as 26 days. The radiologist report indicated a tiny focus of susceptibility in the left superior frontal gyrus (which could be vascular or represent a tiny focus of nonspecific haemosiderin deposition). When analysing T2 relaxometry MRI data for this patient, a significant cluster of voxels was identified in the left superior frontal lobe, with relaxometry times up to 5.28 SD higher than controls (peak relaxometry value = 303.7 ms).

### Case 19

Case study 19 was elbowed in the face during a futsal game and suffered an mTBI as a result. No additional clinical data is available for this patient regarding specific symptom complaints during or post-injury. BIST scores indicate a high rating for 12 different statements, with an initial total score of 117/160. The patient reported an 8/10 for the statement “I don’t like bright lights”, a 9/10 for “I don’t like loud noises”, a 7/10 for “I feel dizzy or like I could be sick”, a 10/10 for “If I close my eyes, I feel like I am at sea”, a 10/10 for “It takes me longer to think”, a 9/10 for “I forget things”, a 9/10 for “I get confused easily”, a 9/10 for “I have trouble concentrating”, an 8/10 for “I get angry or irritated easily”, a 7/10 for “I feel restless”, an 8/10 for “I feel tired during the day”, and an 8/10 for “I need to sleep a lot more or find it hard to sleep at night”. The patient reports no previous mTBIs and the recovery time for the current injury is recorded as 32 days. No significant clusters of voxels were identified when analysing the T2 relaxometry images.

### Case 20

Case study 20 sustained an mTBI while engaging in jiu-jitsu when he was hit with a knee to the head. There are no notes available pertaining to his symptoms at the time of injury, only mention of headaches at his clinical presentation 11 days post-injury. BIST scores were mostly low, however, there was a rating of 6/10 for the statement “I feel tired during the day” and 6/10 for “I need to sleep a lot more or find it hard to sleep at night”, with an initial total BIST score of 28/160. This patient reports two previous mTBIs, with the most recent being in 2012. The recovery time for the current mTBI is reported as 38 days. When analysing the T2 relaxometry images, seven significant voxel clusters were identified. One in the left anterior cingulate cortex, with relaxometry times of up to 17.51 SD above controls (peak relaxometry value = 66.3 ms), one in the left medial frontal lobe with relaxometry times up to 5.68 SD above controls (peak relaxometry value = 121.1 ms), one in the left sensorimotor cortex with relaxometry times up to 4.57 SD above controls (peak relaxometry value = 79.7 ms), one in the left precuneous of the parietal lobe with relaxometry times up to 5.49 SD above controls (peak relaxometry value = 39.0 ms) and one in the left temporoparietal junction with relaxometry times up to 5.60 SD above controls (peak relaxometry value = 100.3 ms). Furthermore, there was a significant cluster in the right anterior cingulate cortex with relaxometry times up to 15.48 SD higher than controls (peak relaxometry value = 73.5 ms) and lastly, in the right superior parietal lobe with relaxometry times up to 4.89 SD higher than controls (peak relaxometry value = 22.1 ms).

## Notes

### Competing Interest Statement

The authors have declared no competing interest.

### Funding Statement

This study was funded by the Health Research Council of New Zealand (HRC)

### Author Declarations

The Health and Disability Ethics Committee of New Zealand gave ethical approval for this work (HDEC 2022 EXP 11078)

